# Awake prone positioning and oxygen therapy in patients with COVID-19: The APRONOX study

**DOI:** 10.1101/2021.01.27.21250631

**Authors:** Orlando R. Perez-Nieto, Diego Escarraman-Martinez, Manuel A. Guerrero-Gutierrez, Eder I. Zamarron-Lopez, Javier Mancilla-Galindo, Ashuin Kammar-García, Miguel A. Martinez-Camacho, Ernesto Deloya-Tomás, Jesús S. Sanchez-Diaz, Luis A. Macías-García, Raúl Soriano-Orozco, Gabriel Cruz-Sánchez, José D. Salmeron-Gonzalez, Marco A. Toledo-Rivera, Ivette Mata-Maqueda, Luis A. Morgado-Villaseñor, Jenner J. Martinez-Mazariegos, Raymundo Flores Ramirez, Josue L. Medina-Estrada, Silvio A. ñamendys-Silva, on behalf of the APRONOX group

## Abstract

**Purpose:** The awake prone position (PP) strategy for patients with acute respiratory distress syndrome (ARDS) is a safe, simple, and cost-effective technique used to improve hypoxemia. We aimed to evaluate the relationship between awake PP (AP) and endotracheal intubation in patients with coronavirus disease (COVID-19).

**Methods:** In this retrospective, multicentre observational study conducted between 1 May and 12 June 2020 in 27 hospitals in Mexico and Ecuador, non-intubated patients with COVID-19 managed with AP or awake supine positioning (AS) were included to evaluate intubation and mortality risk in AP patients through logistic regression models; multivariable adjustment, propensity score analyses, and E-values were calculated to limit confounding. A CART model with cross-validation was also built. This study was registered at https://clinicaltrials.gov/ct2/show/NCT04407468

**Results:** 827 non-intubated patients with COVID-19 in the AP (n=505) and AS (n=322) groups were included for analysis. Less patients in the AP group required endotracheal intubation (23.6% vs 40.4%) or died (20% vs 37.9%). AP was a protective factor for intubation even after multivariable adjustment (OR=0.39, 95%CI:0.28-0.56, p<0.0001, E-value=2.01), which prevailed after propensity score analysis (OR=0.32, 95%CI:0.21-0.49, p<0.0001, E-value=2.21), and mortality (adjusted OR=0.38, 95%CI:0.25-0.57, p<0.0001, E-value=1.98). The main variables associated with PP failure in AP patients were age, lower SpO_2_/FiO_2_, and management with a non-rebreather mask. In the CART model, only two variables were used: SpO_2_/FiO_2_ (F 97.7, p<0.001) and PP (X2 50.5, p<0.001), with an overall percentage of 75.2%.

**Conclusion:** PP in awake hospitalised patients with COVID-19 is associated with a lower risk of intubation and mortality.

## INTRODUCTION

The prone position (PP) in awake, non-intubated patients with acute hypoxemic respiratory failure results in improved oxygenation, as demonstrated by an increase in arterial partial pressure of oxygen (PaO_2_), peripheral arterial oxygen saturation (SpO_2_), and PaO_2_/inspired oxygen fraction (PaO_2_/FiO_2_), without deleterious effects on the level of partial arterial pressure of carbon dioxide (PaCO_2_), pH, respiratory rate (RR), or haemodynamics (1,2). PP combined with non-invasive ventilation (NIV) or high-flow nasal cannula (HFNC) in patients with moderate to severe acute respiratory distress syndrome (ARDS) has been shown to be safe and effective and may prevent intubation (3,4). The pathophysiological mechanism by which PP is useful for ARDS is by increasing functional residual capacity, reducing dead space, reducing intrapulmonary shunts, increasing ventilation in areas dependent of gravity, and relieving the weight that the heart exerts over the lungs (5).

The coronavirus disease (COVID-19) pandemic has unleashed a high global demand for respiratory support, a reason why PP in awake non-intubated patients has become popular and clinical interest has rapidly increased. An early strategy combining PP together with NIV or HFNC has been reported to be associated with reduced intubation and mortality and improved oxygenation (6–8). One further advantage of PP without intubation is that it allows patients to interact with their family during hospitalisation, thereby favouring humanisation of healthcare (9). Nonetheless, few observational studies have included control groups (i.e. awake supine patients managed with NIV or HFNC) and have had conflicting findings. While Ferando et al. (10) and Padrão et al. (11) found no differences in intubation risk between prone and supine patients, Jagan et al. (12) found a reduction in intubation risk for PP patients. Thus, the utility of awake PP remains to be further elucidated in larger observational or randomised studies.

In this multicentre retrospective observational study, we sought to explore the relationship between awake PP and the need for orotracheal intubation, and to develop a model to predict this outcome. The secondary objective was to compare and explore the association between awake PP and mortality risk in the APRONOX study.

## METHODS

### Study design

A multicentre retrospective cohort study was conducted with patients diagnosed with COVID-19 admitted to the emergency department in 27 hospitals in Mexico and Ecuador (Appendix 2). The study was approved by the Health Services Research Committee of the State of Querétaro (registration number 1178/SESEQ-HGSJR/08-05-20) and all other participating centres. This study was prospectively registered in ClinicalTrials.gov (NCT04407468); STROBE recommendations were followed during the reporting of this study.

### Study population and data collection

In each participating hospital centre, data collection was carried out by medical specialists in emergency medicine, respiratory medicine, anaesthesiology, and intensive care medicine, who collected information from patients’ medical records. A separate group of physicians were appointed to review the data obtained and check for plausibility. In cases of doubt physicians in charge at each hospital centre were contacted.

Patients were deidentified by assigning them a code. All patients admitted to the emergency department during the period between 1 May and 12 June 2020 who met the following criteria were ultimately included in the study: 1. Age ≥18 years; 2. Positive SARS-CoV-2 diagnosis; 3. Full inpatient stay at the centre until final outcome; 4. Full clinical records in accordance with the official Mexican standard NOM-004-SSA3-2012 (http://dof.gob.mx/nota_detalle_popup.php?codigo=5272787); and 5. Partial oxygen saturation (SpO_2_) < 94% at room-air partial fraction of inspired oxygen (FiO_2_) upon admission to the emergency department.

Due to the differences in funding and infrastructure between centres, two criteria were employed to standardise SARS-CoV-2 diagnosis: 1. A positive RT-PCR test from a respiratory tract sample; and 2. Positive chest computed tomography (CT) scan with a COVID-19 Reporting and

Data System (CO-RADS) score ≥ 3 (Appendix 3) (13), together with two or more of the following symptoms: eye pain, cough, fever, dyspnoea, headache, arthralgia, or odynophagia. Patients who self-discharged or who were referred to another hospital centre prior to outcome ascertainment, and those with incomplete clinical records, were excluded from the study.

Data recorded were demographic (age, sex) and clinical variables including comorbidities (diabetes, systemic arterial hypertension, obesity, heart disease, lung disease, cancer, liver disease, chronic kidney disease), pre-prone SpO_2_/FiO_2_ ratio (14), supplemental oxygen delivery device used, need for orotracheal intubation, and lethal outcome. FiO_2_ was calculated based on the type of supplemental oxygen delivery device employed: low-flow nasal cannula, high-flow nasal cannula or non-rebreather mask (Appendix 4) (15).

The decision to place patients in the prone position and perform orotracheal intubation was based on individualised medical criteria and was not priorly defined or standardised. The objective of this study was to explore the relationship between orotracheal intubation as the dependent variable and the prone position in awake patients diagnosed with SARS-CoV-2 as an independent variable.

Due to the observational nature of the study and the fact it posed no risk to study participants, convenience sampling was employed with the goal of recruiting the largest number of participants to maximise statistical power.

### Statistical analysis

The clinical and demographic characteristics of the patients were examined for all patients and for those in the awake PP (AP) or awake supine position (AS) groups. Descriptive results for quantitative variables are presented as mean with standard deviation (SD), and frequencies with percentage (%) for qualitative variables. Asymmetry and kurtosis were calculated for quantitative variables. Quantitative comparisons were performed with the independent-samples t-test; qualitative comparisons were done with chi-squared or Fisher’s exact test. Baseline and post-AP SpO_2_/FiO_2_ ratios were compared with the dependent-samples t-test.

To reduce the risk of bias due to unbalanced groups, propensity score analysis was performed through a logistic regression model adjusted for age, sex, the presence of 3 or more comorbidities and baseline SpO_2_/FiO_2_ ratio. Patients were matched in a 1:1 ratio according to the nearest-neighbour matching algorithm; changes in density functions are shown in Appendix 5. All inferential analyses were performed for all patients in the original cohort and for the propensity score-matched cohorts.

Distinct multivariable logistic regression analyses were performed to determine the risk of orotracheal intubation and mortality associated with AP. Variables included in the models were selected by the Enter method; adjustment variables were those which had a p value <0.1 in univariate analyses which have been reported to be associated with higher risk for adverse events (age, sex [men], diabetes, systemic arterial hypertension, obesity, heart disease, cancer, chronic kidney disease), pre-prone SpO_2_/FiO_2_ ratio, and supplemental oxygen delivery device). A multivariable logistic regression model was subsequently created for AP patients to determine the risk of failure to pronation; the variables included in this model were selected with the Stepwise Forward method, including those with a p<0.1 in the final model. Odds ratios (OR) with their 95% confidence interval (95%CI) were calculated. The goodness of fit of the final model was evaluated with the Hosmer-Loemeshow statistic, and the discrimination of the model was determined by calculating the area under the curve (AUC). The risk of failure to AP according to age and baseline SpO_2_/FiO_2_ ratio were graphed through the smoothing spline method. E-values for the lower bound of the confidence intervals were calculated to determine the value at which an unmeasured confounding factor could potentially alter the observed effect of AP on the outcomes and drive them to a non-significant value (16). Regression analyses were verified through residual analysis.

A classification and regression tree (CART) model were constructed with cross-validation, the QUEST (Quick, Unbiased, Efficient, Statistical Tree) growing method, and pruning control to prevent overfitting (reduce standard error). This methodology is based on developing hierarchical binary classification trees with sensitivity analysis based on the Gini index (17).

A systematic review of studies of AP was conducted; the search strategy and inclusion criteria for studies are provided in Appendix 6. Results of eligible studies were summarised alongside the propensity score-matched cohort of APRONOX in a forest plot and the overall risk for intubation for patients in AP vs AS was calculated.

Missing values were not imputed. A p-value <0.05 was used to define bilateral statistical significance. All analyses and graphs were created with the SPSS software v.21, R software v.3.4.2, and RevMan 5.3.

## RESULTS

Out of 932 patients identified across all 27 hospital centres, 827 patients were ultimately included for analysis (Figure 1). Descriptive results for all patients are provided in Table 1. Among all 927 patients, 227 (27.4%) were female and mean age was 54.3 (SD:14.2) years, with most patients being in the 50 to 59-year category (25.3%). The most prevalent comorbidities were diabetes (38.1%) and hypertension (34.5%). Most patients were managed with low-flow nasal cannulas (48.6%). The characteristics of patients in the AP and AS groups, in both the unmatched and matched cohorts, are provided in Table 2. A lesser proportion of patients in the AP group required endotracheal intubation (23.6% vs 40.4%) or had a lethal outcome (20% vs 37.9%). After propensity score matching, these differences prevailed. The SpO_2_/FiO_2_ ratio in the AP group was statistically significantly higher after PP (217.42, SD: 81.9) compared with baseline values (182.39, SD: 81.91), with a mean difference of 35.03 (95%CI: 29.99-40.06, p<0.0001) units.

**Table 1.** Descriptive characteristics of patients in the APRONOX cohort

|  |  |
| --- | --- |
| **Demographic variables** |  |
| Age, years | 54.3 (14.2) |
| Age categories, n (%) |  |
| <20 | 1 (0.1) |
| 20-29 | 29 (3.5) |
| 30-39 | 101 (12.2) |
| 40-49 | 194 (23.5) |
| 50-59 | 209 (25.3) |
| 60-69 | 162 (19.6) |
| ≥70, n | 131 (15.8) |
| Sex |  |
| Women, n (%) | 227 (27.4) |
| Men, n (%) | 600 (72.6) |
| **Clinical variables** |  |
| Diabetes, n (%) | 315 (38.1) |
| Systemic arterial hypertension, n (%) | 285 (34.5) |
| Obesity, n (%) | 119 (14.4) |
| Heart disease, n (%) | 17 (2.1) |
| Lung disease, n (%) | 41 (5) |
| Cancer, n (%) | 10 (1.2) |
| Liver disease, n (%) | 5 (0.6) |
| Chronic kidney disease, n (%) | 35 (4.2) |
| **Baseline SpO<sub>2</sub>/FiO<sub>2</sub> ratio** | 189.5 (81.6) |
| **Awake prone, n (%)** | 505 (61.1) |
| **Not awake prone, n (%)** | 322 (38.9) |
| **Supplemental oxygen delivery device** |  |
| Low-flow nasal cannula, n (%) | 402 (48.6) |
| High-flow nasal cannula, n (%) | 83 (10) |
| Non-rebreather mask, n (%) | 342 (41.4) |
| **Adverse events, n (%)** |  |
| Intubation, n (%) | 249 (30.1) |
| Mortality, n (%) | 220 (26.6) |
| Failure to the prone, n (%) | 119 (23.6)* |
| *Percentage calculated out of all awake prone-positioned patients. |  |
| FiO<sub>2</sub>: Inspired oxygen fraction; SpO<sub>2</sub>: peripheral arterial oxygen saturation. |  |

**Table 2.** Comparison of demographic, clinical, and outcome characteristics of patients in the awake prone and awake supine groups in both the unmatched and propensity score-matched cohorts.

|  | **Unmatched** |  |  | **Matched** |  |  |
| --- | --- | --- | --- | --- | --- | --- |
|  | **Awake**<br>**supine**<br>**(n =**<br>**322)** | **Awake**<br>**prone**<br>**(n =**<br>**505)** | **p-value** | **Awake**<br>**supine**<br>**(n =**<br>**306)** | **Awake**<br>**prone**<br>**(n = 306)** | **p-value** |
| **Demographic**<br>**variables** |  |  |  |  |  |  |
| Age, years | 55.8<br>(14.5) | 53.4<br>(13.9) | 0.02 | 55.8<br>(14.6) | 54.9 (14.4) | 0.5 |
| Women | 92 (28.6) | 135<br>(26.7) | 0.6 | 85 (27.8) | 78 (25.5) | 0.5 |
| Men | 230<br>(71.4) | 370<br>(73.3) |  | 221<br>(72.2) | 228 (74.5) |  |
| **Clinical variables** |  |  |  |  |  |  |
| Diabetes | 121<br>(37.6) | 194<br>(38.4) | 0.8 | 115<br>(37.6) | 114 (37.3) | 0.9 |
| Systemic arterial<br>hypertension | 119 (37) | 166<br>(32.9) | 0.2 | 110<br>(35.9) | 107 (35) | 0.8 |
| Obesity | 45 (14) | 74 (14.7) | 0.8 | 45 (14.7) | 43 (14.1) | 0.8 |
| Heart disease | 4 (1.2) | 13 (2.6) | 0.2 | 3 (1.0) | 11 (3.6) | 0.06 |
| Lung disease | 17 (5.3) | 24 (4.8) | 0.7 | 16 (5.2) | 19 (6.2) | 0.6 |
| Cancer | 8 (2.5) | 2 (0.4) | 0.02 | 7 (2.3) | 1 (0.3) | 0.07 |
| Liver disease | 3 (0.9) | 2 (0.4) | 0.4 | 3 (1) | 2 (0.7) | 0.9 |
| Chronic kidney<br>disease | 12 (3.7) | 23 (4.6) | 0.6 | 12 (3.9) | 14 (4.6) | 0.8 |
| SpO<sub>2</sub>/FiO<sub>2</sub> ratio | 201.1<br>(89.8) | 182.4<br>(75.4) | 0.002 | 202.6<br>(89.6) | 198.4 (82.9) | 0.5 |
| **Supplemental**<br>**oxygen delivery**<br>**device** |  |  |  |  |  |  |
| Low-flow nasal<br>cannula | 149<br>(46.3) | 253<br>(50.1) | 0.3 | 140<br>(45.8) | 157 (51.3) | 0.2 |
| High-flow nasal<br>cannula | 22 (6.8) | 61 (12.1) | 0.01 | 22 (7.2) | 37 (12.1) | 0.04 |
| Non-rebreather mask | 151<br>(46.9) | 190<br>(37.6) | 0.008 | 144<br>(47.1) | 111 (36.3) | 0.007 |

| **Adverse events** |  |  |  |  |  |  |
| --- | --- | --- | --- | --- | --- | --- |
| Intubation | 130<br>(40.4) | 119<br>(23.6) | <0.0001 | 118<br>(38.6) | 65 (21.2) | <0.0001 |
| Mortality | 120<br>(37.9) | 100 (20) | <0.0001 | 113<br>(36.9) | 59 (19.3) | <0.0001 |

FiO<sub>2</sub>: Inspired oxygen fraction; SpO<sub>2</sub>: peripheral arterial oxygen saturation.

**Figure 1.**
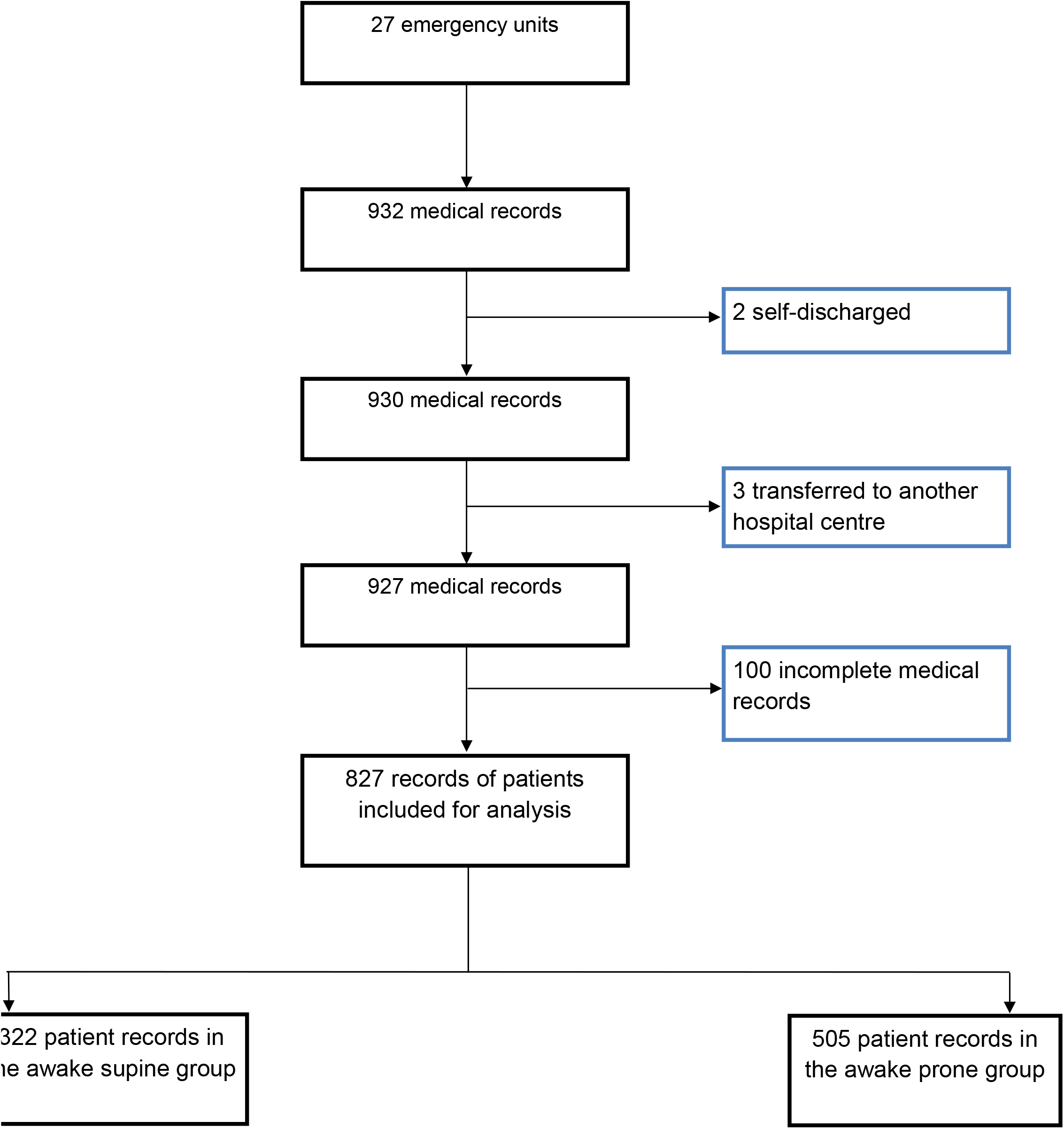
Flow diagram of participants included in the APRONOX cohort

The results of univariable logistic regression models for orotracheal intubation risk are provided in Table 3, for both the unmatched and matched cohorts. The main risk factors identified were age, diabetes, arterial hypertension, obesity, heart disease, cancer, a baseline SpO_2_/FiO_2_ <100 or between 100 and 199, and management with a non-rebreather mask. Ventilation in AP was a protective factor for orotracheal intubation even after multivariable adjustment Table 4 for confounding variables (Adjusted OR=0.39, 95%CI:0.28-0.56, p<0.0001, E-value=2.01), which prevailed after propensity score analysis (Adjusted OR=0.32, 95%CI:0.21-0.49, p<0.0001, E-value=2.21). Similarly, ventilation in AP was a protective factor for mortality (Adjusted OR=0.38, 95%CI:0.25-0.57, p<0.0001, E-value=1.98, Goodness of fit: Hosmer-Lemeshow X2=11.7, p=0.1 AUC=0.80, 95%CI:0.77-0.84, p<0.0001) even after multivariable adjustment in propensity score analyses.

**Table 3.** Results of univariable logistic regression analyses of orotracheal intubation risk in patients with awake prone positioning.

|  | **Unmatched** |  | **Matched** |  |
| --- | --- | --- | --- | --- |
|  | **OR (95% CI)** | **p value** | **OR (95% CI)** | **p value** |
| Awake prone | 0.46 (0.34-0.62) | <0.0001 | 0.43 (0.30-0.62) | <0.0001 |
| **Demographic variables** |  |  |  |  |
| Age, years | 1.02 (1.004-1.03) | 0.007 | 1.01 (1.001-1.03) | 0.03 |
| Sex (Men) | 0.91 (0.70-1.37) | 0.9 | 1.07 (0.72-1.59) | 0.7 |
| **Clinical variables** |  |  |  |  |
| Diabetes | 1.70 (1.26-2.30) | 0.001 | 1.71 (1.21-2.44) | 0.003 |
| Systemic arterial hypertension | 1.61 (1.19-2.19) | 0.002 | 1.60 (1.12-2.29) | 0.009 |
| Obesity | 2.01 (1.35-2.99) | 0.001 | 2.48 (1.57-3.93) | <0.0001 |
| Heart disease | 3.41 (1.28-9.07) | 0.01 | 4.39 (1.45-13.27) | 0.009 |
| Lung disease | 1.36 (0.71-2.62) | 0.4 | 1.08 (0.52-2.25) | 0.8 |
| Cancer | 9.56 (2.02-45.35) | 0.004 | 17.02 (2.08-139.38) | 0.008 |
| Liver disease | 3.51 (0.58-21.15) | 0.2 | 3.56 (0.59-21.48) | 0.2 |
| Chronic kidney disease | 1.39 (0.69-2.81) | 0.4 | 1.77 (0.79-3.92) | 0.2 |
| **Baseline SpO<sub>2</sub>/FiO<sub>2</sub> ratio** |  |  |  |  |
| <100 | 5.69 (3.48-9.31) | <0.0001 | 6.98 (3.86-12.64) | <0.0001 |
| 100-199 | 3.69 (2.57-5.29) | <0.0001 | 4.08 (2.72-6.12) | <0.0001 |
| ≥200 | Reference |  | Reference |  |
| **Supplemental oxygen delivery device** |  |  |  |  |
| Low-flow nasal cannula | 0.27 (0.19-0.38) | <0.0001 | 0.24 (0.16-0.35) | <0.0001 |
| High-flow nasal cannula | 0.77 (0.46-1.29) | 0.3 | 0.78 (0.42-1.44) | 0.4 |
| Non-rebreather mask | 3.94 (2.88-5.39) | <0.0001 | 4.45 (3.08-6.44) | <0.0001 |

95%CI: 95% confidence interval; FiO<sub>2</sub>: Inspired oxygen fraction; OR: odds ratio; SpO<sub>2</sub>: peripheral arterial oxygen saturation.

**Table 4.** Results of multivariable logistic regression analyses of orotracheal intubation risk in patients with awake prone positioning, adjusted by confounding variables.

|  | **Unmatched\*** |  | **Matched\*\*** |  |
| --- | --- | --- | --- | --- |
|  | **OR (95% CI)** | **P value** | **OR (95% CI)** | **P value** |
| Awake prone | 0.39 (0.28-0.56) | <0.0001 | 0.38 (0.25-0.58) | <0.0001 |
| Age, years | 1.01 (0.99-1.02) | 0.3 | 1.01 (0.99-1.02) | 0.4 |
| Sex (Men) | 1.22 (0.82-1.81) | 0.3 | 1.26 (0.79-2.03) | 0.3 |
| Diabetes | 1.46 (1.01-2.10) | 0.04 | 1.38 (0.89-2.13) | 0.2 |
| Systemic arterial hypertension | 1.24 (0.85-1.81) | 0.3 | 1.15 (0.74-1.79) | 0.5 |
| Obesity | 1.37 (0.85-2.21) | 0.2 | 1.45 (0.82-2.57) | 0.2 |
| Heart disease | 6.42 (2.06-20.02) | 0.001 | 11.31 (3.05-41.88) | <0.0001 |
| Cancer | 6.61 (0.93-49.99) | 0.06 | 10.95 (0.91-132.13) | 0.06 |
| Chronic kidney disease | 1.15 (0.47-2.79) | 0.8 | 1.50 (0.53-4.25) | 0.4 |
| Baseline SpO<sub>2</sub>/FiO<sub>2</sub> ratio | 0.99 (0.98-0.99) | <0.0001 | 0.98 (0.98-0.99) | <0.0001 |
| Low-flow nasal cannula | 1 | - | 1 | - |
| High-flow nasal cannula | 0.94 (0.50-1.74) | 0.8 | 1.12 (0.53-2.36) | 0.8 |
| Non-rebreather mask | 2.56 (1.73-3.77) | <0.0001 | 2.92 (1.82-4.69) | <0.0001 |
| *Goodness of fit: Hosmer-Lemeshow $X^2=12.43$, $p=0.1$; AUC=0.76, 95%CI:0.72-0.79, $p<0.0001$.<br>**Goodness of fit: Hosmer-Lemeshow $X^2=2.43$, $p=0.9$; AUC=0.82, 95%CI:0.78-0.85, $p<0.0001$.<br>95%CI: 95% confidence interval; AUC: area under de curve; FiO<sub>2</sub>: Inspired oxygen fraction; OR: odds ratio; SpO<sub>2</sub>: peripheral arterial oxygen saturation. |  |  |  |  |

The CART model is shown in Figure 2 and the main characteristics are broken down in Appendix 7. Of all variables, only two were statistically significant, in hierarchical order: SpO_2_/FiO_2_ (F 97.7, p = 0.00) and PP (X^2^ 50.5, p = 0.00), with an overall percentage of 75.2%.

**Figure 2.**
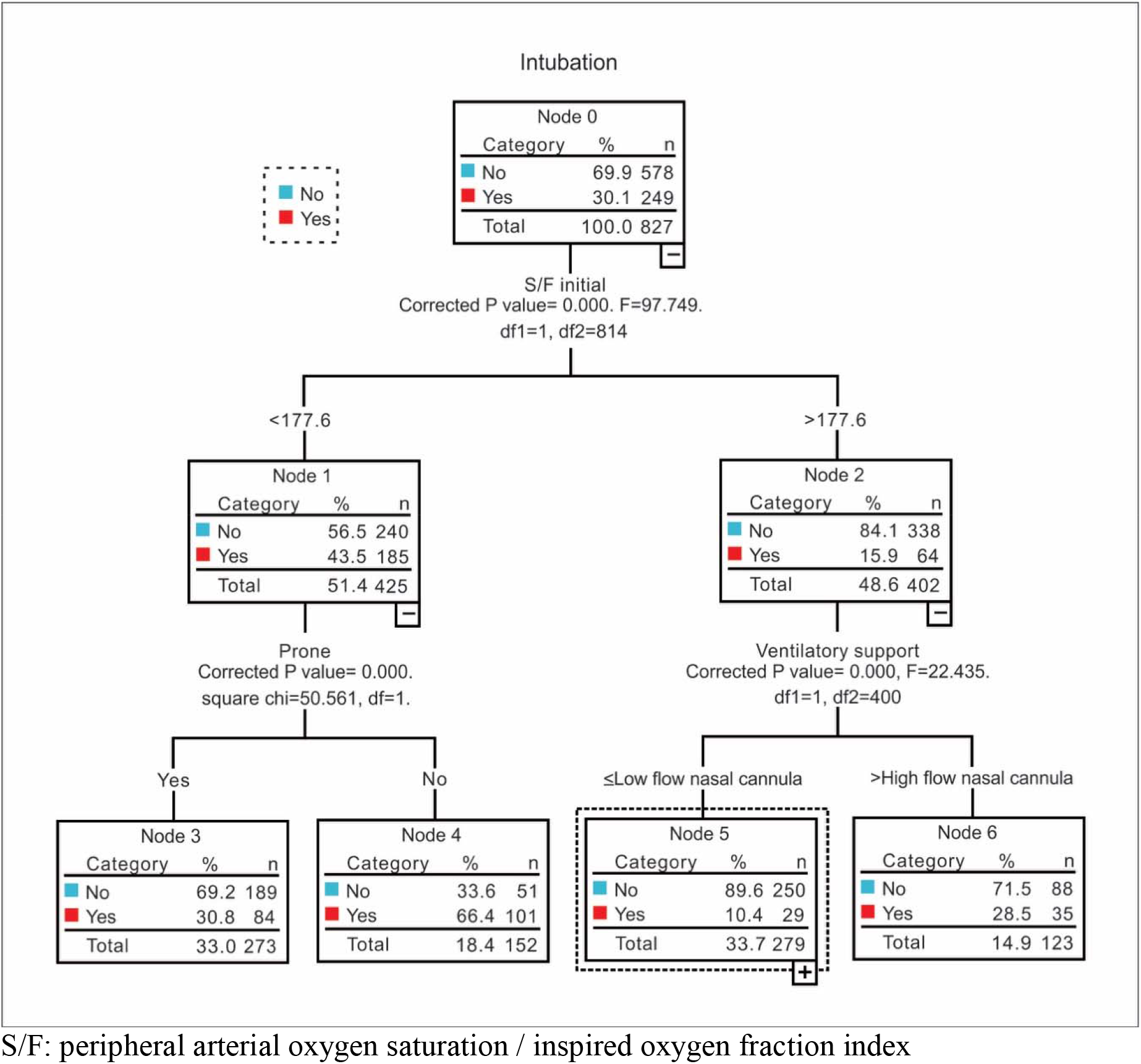
Classification and regression tree (CART) model for the APRONOX study

The main variables associated with PP failure in AP patients were age (OR=1.02, 95%CI: 1.35-5.72, p=0.005), SpO_2_/FiO_2_ <100 (OR=2.78, 95%CI: 1.35-5.72, p=0.005), SpO_2_/FiO_2_ 100-199 (OR=2.18, 95%CI: 1.31-3.64, p=0.003), and management with a non-rebreather mask (OR=2.17, 95%CI: 1.34-3.49, p=0.002), Goodness of fit: Hosmer-Lemeshow X2=10.52, p=0.2; AUC=0.70, 95%CI:0.64-0.74, p<0.0001. The distribution of risk for increases in age and baseline SpO_2_/FiO_2_ are shown in Figure 3.

**Figure 3.**
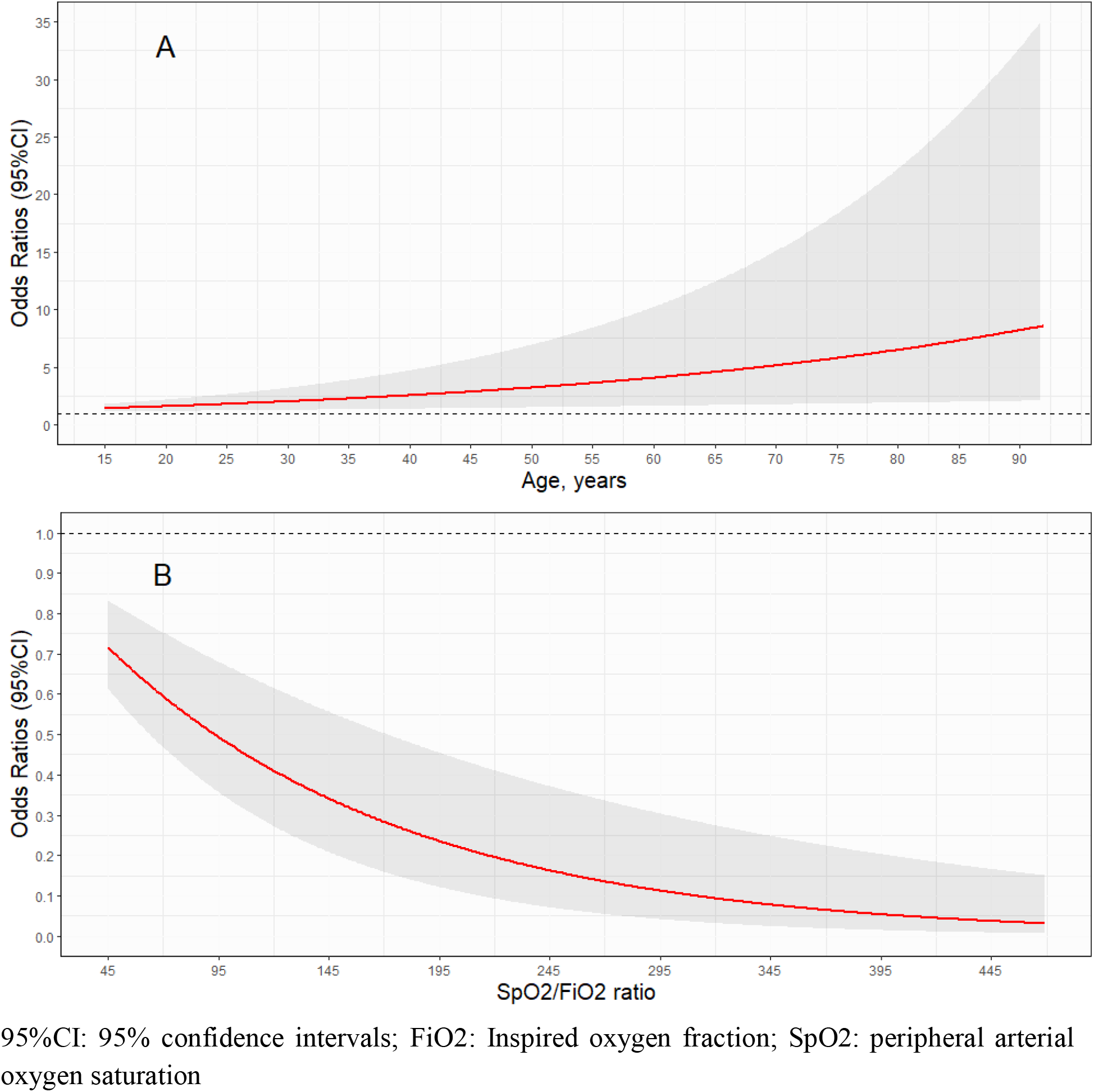
Risk of prone positioning failure according to age and baseline SpO_2_/FiO_2_

After the search of the literature, 54 records were obtained, of which only 3 studies were comparison-group studies including both AP and AS patients for which sufficient information was available to the overall risk for intubation, which are summarised alongside the APRONOX study in Figure 4.

**Figure 4.**
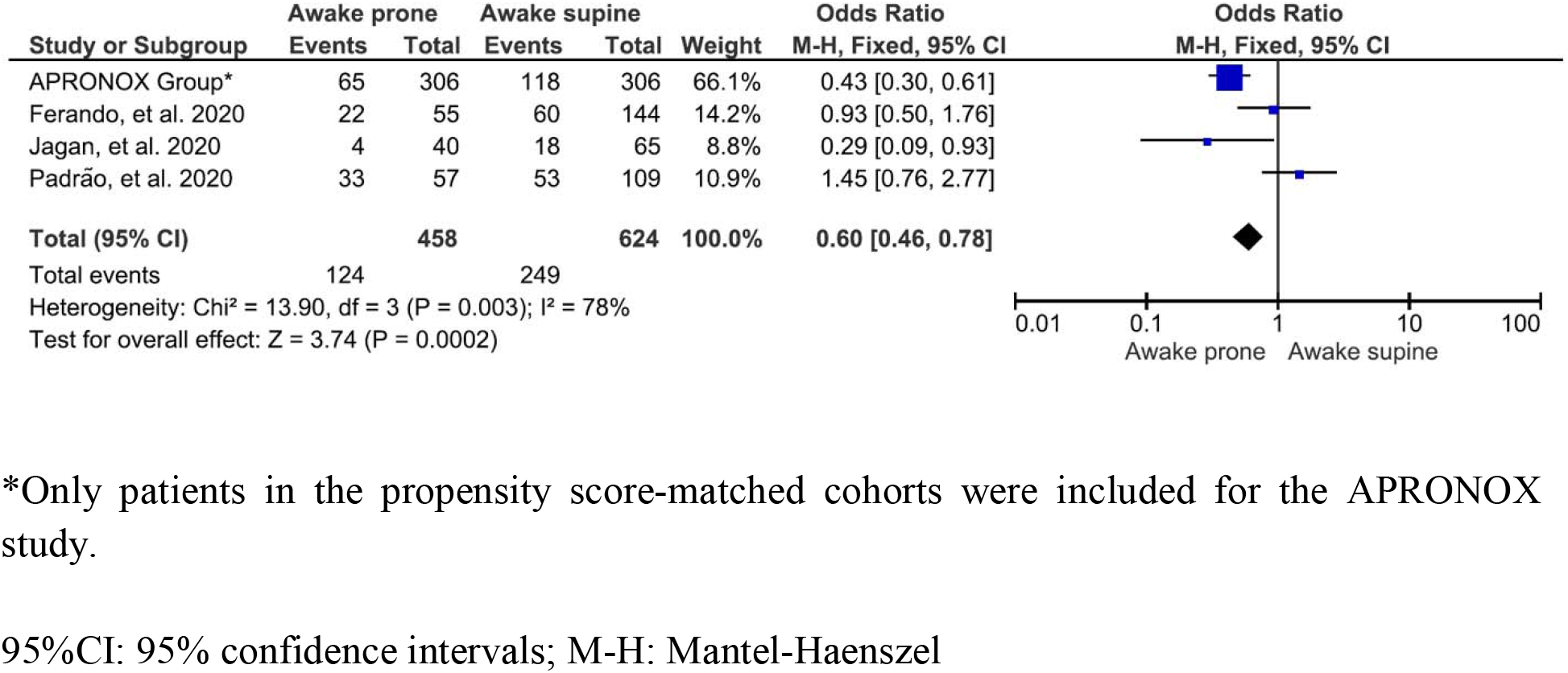
Forest plot of overall risk of orotracheal intubation in studies retrieved by the search strategy and the APRONOX cohort.

## DISCUSSION

In this multicentre observational study, we aimed to evaluate the association between awake prone positioning and orotracheal intubation, as well as predictors of failure to prone positioning and mortality in hospitalised patients with COVID-19. Even after multivariable adjustment and propensity score analyses, prone positioning in non-intubated patients was associated with lower intubation and mortality risk. We further developed a CART model to evaluate the relationship between AP and intubation.

Patients in our cohort were younger (mean age 53.4 years) than those in other studies (56.0-65.8) (10–12); hospitalised patients with COVID-19 in Mexico have been reported to be young (18). The prevalence of comorbidities in our study is similar to that reported in a population-based sample of Mexican patients hospitalised with COVID-19, although diabetes was more common in our study (38.1% vs 29.2%), whereas obesity (14.4% vs 22.5%) and heart disease (2.1% vs 4.4%) were less frequent (18).

The overall intubation rate in the APRONOX cohort was higher (30.1%) than that reported for hospitalised patients with COVID-19 in Mexico City (20.2%) (18); however, limited access to beds with ventilators in Mexico has been reported (19). Intubation rates for patients in the unmatched AP (23.6%) and AS (40.4%) cohorts fall within those reported in previous studies (10–58% and 27.7–49%, respectively) (10–12). Awake prone positioning in our study was associated with decreased intubation risk even after multivariable adjustment in both the unmatched and propensity-score matched cohorts, with an E-value of 2.01 and 2.21, respectively, which reflects that in order to drive this association to be non-significant, an unmeasured risk factor should have a lower-limit confidence interval that at least doubles the risk of the outcome between both groups. Out of all comorbidities, only diabetes and heart disease were associated with increased intubation risk after multivariable adjustment, although diabetes was no longer a risk factor after propensity score analysis. Baseline SpO_2_/FiO_2_ was associated with reduced intubation risk. The mortality rate reported in our study was 19.8%, comparable to 23.4% (12) and 27% (10) in other studies.

Regarding variables associated to failure to awake prone positioning, age, SpO_2_/FiO_2_, and the use of a non-rebreather mask were the main variables associated. The distribution of risk for quantitative values of age show that the risk of failure is higher with increasing ages, whereas higher baseline SpO_2_/FiO_2_ have the lowest failure risks.

The decision rules obtained from the CART model were as follows:

1. Regardless of sex, patients with an initial SpO_2_/FiO_2_ ratio ≤177.6 have a 43.5% chance of being intubated, while patients with an initial SpO_2_/FiO_2_ ratio >177.6 have only a 15.9% chance of being intubated.
2. Among patients with an initial SpO_2_/FiO_2_ ratio ≤177.6, when prone positioning is not used, the likelihood of intubation increases to 66.4%, while PP reduces this figure to 30.8%.

Thus, when the SpO_2_/FiO_2_ ratio upon admission is under 177.6, risk of intubation is increased. This is comparable to findings by Thompson et al., who found that the mean difference in the intubation rate between patients with SpO_2_ at 95% or greater and patients with SpO_2_ less than 95% one hour after initiation of PP was 46% (8).

PP has been presented as one the most cost-effective strategies to treat patients with COVID-19. In countries with limited oxygen delivery devices, and a shortage of ventilators, AP has been routinely used to avoid intubating patients with COVID-19 (20). Nonetheless, conflicting evidence from observational studies for AP exists.

The supine position alters pulmonary function in patients with respiratory insufficiency due to the gravitational differences between dependent and non-dependent regions, resulting in a more negative pleural pressure (Ppl), increasing transpulmonary pressure (TPP) in non-dependent areas (more distension), and producing the opposite effect in dependent areas where Ppl is less negative and TPP is lower (less distension). Ventilation in the PP causes even distribution of TPP, favouring uniform ventilation (21). Approximately 45 years ago, PP was shown to increase oxygenation in patients with respiratory insufficiency, primarily by improving the ventilation-perfusion ratio (V/Q) (22).

PP has been evaluated in hospitalised patients with respiratory failure due to COVID-19, having observed improvements in SpO_2_ and PaO_2_, decreased respiratory rate (RR), decreased need for intubation and possible reductions in mortality, in addition to being cost-free (8,23–25). As summarised in Figure 4, only three other studies to date have evaluated intubation risk among AP compared with AS. While Ferando et al. and Padrão et al. found no differences in intubation risk, Jagan et al found reduced intubation risk in AP patients (10–12). The APRONOX study is the largest study to date evaluating the effect of AP on intubation risk.

Regarding modality of ventilation, the use of a non-rebreather mask was associated with greater risk of intubation and failure to prone positioning, whereas other modalities of ventilation were not. There is documented evidence of the correlation between the oxygen saturation/fraction of inspired oxygen (SaO_2_/FiO_2_) ratio and the partial pressure of oxygen/fraction of inspired oxygen (PaO_2_/FiO_2_) ratio, with the advantage that the SaO_2_/FiO_2_ ratio only relies on a pulse oximeter, with no need to perform a blood gas test, thereby highlighting the value of validated cost-effective strategies (14).

Unsupervised machine learning algorithms are being increasingly used in medicine as techniques to support the development of models to improve clinical decisions. Such models have been used in attempts to achieve early, adequate predictions of ARDS with the goal of improving diagnosis, treatment and monitoring, using clinical and demographic data (26), or by adding genomic information to evaluate response to treatment (27) or to identify different phenotypes of a disease (28). Our model contributes valuable information that may be used to decide whether to initiate PP in a patient or not. Although no statistically significant association was found between types of O_2_ delivery device, it was shown that PP determines whether intubation is needed or not, regardless of the type of device used. These prediction techniques could serve as a guide for healthcare workers in resource-strained settings to guide decision making (29).

Our study has the following limitations: 1) O_2_ delivery devices were not standardised to a unique device; 2) the number of hours of PP varied between hospitals and patients; and 3) no precise criteria were established to consider intubation in patients requiring IMV. Nonetheless, this reflects how PP is used in real-world settings. The strengths of our research include: 1) this is the largest study evaluating AP to date; 2) the large number of hospitals included; and 3) the fact that various O_2_ delivery devices were employed, showing that the benefits of PP are not necessarily unique to NIV or HFNC devices, which are costlier and not always available.

PP in spontaneously breathing patients with acute hypoxemic respiratory insufficiency may be a justifiable treatment modality, given the improvements in oxygenation and its physiological benefits, but the decision to intubate is based on the clinician’s best judgement and intubation should not be delayed if under consideration. Close clinical evaluation of patients is key to avoid poor outcomes. Studies of PP in non-intubated patients are challenging and randomized controlled trials are warranted to fully elucidate their usefulness since this is an easy to administer, safe, and reproducible intervention (30).

## CONCLUSION

PP in awake hospitalised patients with COVID-19 is associated with a lower risk of intubation and mortality.

## Data Availability

All data that support the findings of this study will be available from the corresponding author upon reasonable request.

## Acknowledgements

**Healthcare workers treating COVID-19 patients:** Edgard Díaz Soto, Jaziel López Pérez, José Antonio Meade Aguilar, Rubén Rodríguez Blanco, José Luis Patiño Pérez, Janisia Rodríguez Solís, Maribel Santosbeña Lagunes, Alberto Calvo Zúñiga, Manuel de Jesús Santaella Sibaja, Luis Iván Contreras Ley, María Alejandra Sicsik Aragón, Yessica Bernal Luna, Carlos Baez Ambriz, Yanira Jiménez Blancas, Alejando Ayala Mata, Tania Gabriela Ramírez Lira, Iván Avalos Flores, Edwing Díaz Rodríguez, Roberto Robles Godínez, Eduardo Espino López, Hugo Francisco Díaz Ramírez, Concepción Mendoza Fragoso, Oliver Garaz Trujillo, and Jesús Elías Paredes Flores.

The APRONOX Group wishes to thank Rubén Rodriguez Blanco, Jose Luis Patiño Pérez, Janisia Rodriguez Solis, Maribel Santosbeña Lagunes, Alberto Calvo Zuñiga, Manuel de Jesus Santaella Sibaja, Luis Iván Contreras Ley, María Alejandra Sicsik Aragon, Yessica Bernal Luna, Carlos Baez Ambriz, Yanira Jimenez Blancas, Alejando Ayala Mata, Tania Gabriela Ramirez Lira, Iván Avalos Flores, Edwing Díaz Rodriguez, Roberto Robles Godinez, Eduardo Espino López, Hugo Francisco Díaz Ramirez, Concepción J. Mendoza Fragoso, Oliver Garaz Trujillo, and Jesús Elias Paredes Flores for their help in providing care to patients with COVID-19.

## Appendix 1. Full list of authors with place of affiliation

### Writing Committee

Orlando Ruben Perez Nieto MD; *Hospital General San Juan del Río, Querétaro. Intensive Care Unit*. Manuel Alberto Guerrero Gutierrez MD; *Instituto Nacional de Cancerología, Mexico City. Intensive Care Unit*. Eder Ivan Zamarron Lopez MD; *Hospital CEMAIN Tampico, Tamaulipas. Intensive Care Unit*. Ernesto Deloya Tomas MD; *Hospital General San Juan del Río, Querétaro. Head of Intensive Care Unit*. Javier Mancilla-Galindo MBBS; *Instituto Nacional de Enfermedades Respiratorias, México City. Respiratory Medicine Fellow*. Ashuin Kammar-García, PhD; *Instituto Nacional de Nutrición y Ciencias Médicas “Salvador Zubiran”, Mexico City. Emergency Department*. Raúl Soriano Orozco; *Hospital de Alta Especialidad T1 IMSS, León, Guanajuato. Intensive Care Unit*. Gabriel Cruz Chavez MD; *Clínica Hospital Mérida ISSSTE. Head of Intensive Care Unit*. Jose David Salmeron Gonzalez MD; *Hospital General “Miguel Silva”, Morelia, Michoacán. Intensive Care Unit*. Marco Antonio Toledo Rivera MD; *Hospital SEDNA, Mexico City. Head of Intensive Care Unit*. Luis Antonio Morgado Villaseñor MD; *Hospital General de Zona IMSS No*.*15 Reynosa, Tamaulipas. Intensive Care Unit*.

Jenner Jose Martínez Mazariegos MD; *Hospital Vida Mejor ISSSTECH Tuxtla Gutiérrez, Chiapas. Intensive Care Unit*. Silvio Antonio Ñamendys Sylva MD, MSc, FCCP, FCCM; *Instituto Nacional de Cancerología and Instituto Nacional de Nutrición y Ciencias Médicas “Salvador Zubiran”, Mexico City. Head of Intensive Care Unit*.

### General Research Committee

Diego Escarraman Martinez MD, MSc; *Centro Médico Nacional IMSS “La Raza”, Mexico City. Department of Anaesthesiology*. Miguel Angel Martinez Camacho, PT, MSc; *Hospital General de Mexico, Mexico City. Intensive Care Unit*. Ivette Mata Maqueda MD, MSc. DSc; *Secretaría de Salud del Estado de Querétaro, Ethics and Research Committee*. Jesús Salvador Sánchez Díaz MD, MSc; *Hospital de Alta Especialidad IMSS “Adolfo Ruiz Cortines” Veracruz, Veracruz. Intensive Care Unit*. Luis Alberto Macias Garcia, MD, MSc; *Hospital Regional ISSSTE “Fernando Quiroz Gutiérrez”, Mexico City. Intensive Care Unit*. Josué Luis Medina Estrada MD; *Hospital Regional No. 1 IMSS “Vicente Guerrero”, Acapulco, Guerrero. Intensive Care Unit*.

### Local researchers

**Hospital SEDNA, Mexico City** *Ivette Zapata Centeno MD, Intensive Care Unit; Cecilia Hernández Fernández MD*. **Hospital General de Zona No. 33 IMSS Bahía de Banderas, Nayarit:** *Francisco Agustín Martínez Ayuso MD, Intensive Care Unit*, **Hospital General “Dr. Miguel Silva”, Morelia, Michoacán:** *José David Salmerón González MD, Intensive Care Unit. Juan Manuel Angeles Uribe MD, Emergency Department*. **Centro Médico Lic. Adolfo López Mateos, ISEM, Toluca, State of Mexico:** *Aaron Alacio Ávila MD, Intensive Care Unit. Abad Quetzalcoatl Ortega Pérez MD, Head of Intensive Care Unit*. **Centro Medico Nacional 20 de Noviembre, ISSSTE, Mexico City:**

*Jessica Selene Cancino Cuevas MD, Intensive Care Unit. Alberto Hilarion de la Vega Bravo MD, Head of Intensive Care Unit*. **Clínica Hospital Mérida ISSSTE, Mérida, Yucatán:** *Gabriel Cruz Sanchez MD. Clínica Hospital Mérida ISSSTE. Head of Intensive Care Unit*. **Hospital General Dr. Enrique Cabrera, Mexico City:** *Ivan Ilescas Martinez MD, Emergency Department. Lilian Saraí Ramirez Serrano MD, Emergency Department*. **Hospital Regional de Alta Especialidad de Zumpango, State of Mexico:**

*Areli Patricia Ortíz Jimenez MD, Emergency Department. María José Pecero Hidalgo MD, Department of Pneumology*. **Hospital Estatal de Atención de Pacientes COVID-19, Leon, Guanajuato:** *Jorge Adalid Díaz Rodriguez MD, Respiratory Care Unit*. **Hospital Juárez de México, Mexico City:** *Juan Carlos Betancourt Aldana Villarruel MD, Department of Cardiology. José Carlos Gasca Aldama MD, Respiratory Care Unit*.

*Ruben Nicolas Mendoza MD, Department of Cardiology. Luis Fausto García Mayen MD, Head of Cardiovascular Medicine Department*. **Hospital General Tuxtepec, Oaxaca:**

*Jesús Ariben Servando Álvarez Ramirez MD, Respiratory Care Unit. Enrique Fleuvier Morales López MD. Respiratory Care Unit*. **Hospital General de San Juan del Río, San Juan del Río, Querétaro**. *Jorge López Fermin MD, Intensive Care Unit, Tania Mondragon Labelle MD, Intensive Care Unit, Gabriela Castillo Gutierrez MD, Intensive Care Unit, Jorge Daniel Carrión Moya MD, Intensive Care Unit, María Guadalupe Olvera Ramos MD. Intensive Care Unit, Manuel Alfredo Díaz Martínez, MD, Department of Anaesthesiology*. **Hospital Santo Tomás Querétaro, Querétaro**. *Cristobal Meneses Olguín MD, Head of Respiratory Care Unit, Andrea de la Torre Ritscher MD, Respiratory Care Unit. Lizbeth Franco Morales MD, Respiratory Care Unit. Martin de Jesus Reyna Ramirez MD, Respiratory Care Unit. Angélica Del Carmen Chimal Ayohua MD, Respiratory Care Unit*. **Hospital General de Zona No. 48 “San Pedro Xalpa”, Mexico City**. *César Daniel Alonso Bello MD, Internal Medicine Department. Edgar Pérez Barragán MD. Department of Infectiology*. **Hospital General de Zona No. 71 Veracruz, Veracruz**. *Oscar Rodrigo Jimenez Flores, Intensive Care Unit. Ulises Espinosa Hernandez MD, Emergency Department*. **Hospital Comunitario de Ocuituco, Morelos:** *Iván Hernández Bernabé MD. Internal Medicine Department*.

*Yuliana Young Peralta MD, Emergency Department. José Ramón Arteaga Solis MD, Medical Director*. **Hospital General Regional No 200 Tecámac, State of Mexico:**

*Josafat Jesús Gutierrez de la Cruz MD. Emergency Department*. **Unidad Médica de Alta Especialidad IMSS No. 189 “Adolfo Ruiz Cortines”:** *Jesús Salvador Sánchez Díaz MD, Intensive Care Unit. Xiomara García Montes MD, Emergency Department*. **Hospital General Regional IMSS No. 251 Metepec, State of Mexico:** *Carlos Mendiola Villalobos MD, Emergency Department. Alejandro Esquivel Loza MD, Internal Medicine Department*. **Hospital General Regional ISSSTE “Fernando Quiroz Gutiérrez”, Mexico City:** *María Concepción Gonzalez Belmont MD*, **Hospital General de Querétaro, Querétaro:** *Raul Arturo Gonzalez Toribio MD, Intensive Care Unit*.

*Alicia Alejandra Rico Pérez MD, Emergency Department. ArjunaAliel Sotomayor Zavala MD, Emergency Departmen*. **Hospital IESS “Manuel Ygnacio Monteros”, Loja, Ecuador:** *Tatiana Maribel Merino Mijas MD, Intensive Care Unit. Maria Eugenia Abad Guarnizo MD. Intensive Care Unit*. **Hospital Materno de Celaya, Guanajuato:**

*Karen Pamela Pozos Cortes MD, Head of Intensive Care Unit*. **Hospital CEMAIN, Tampico, Tamaulipas:** *María Angelica Sánchez Cepeda, MD, Head of Intensive Care Unit*. **Hospital Star Medica Luna Parc, Cuautitlán Izcalli, State of Mexico:**

*Marco Antonio Villagrana Rodríguez. Intensive Care Unit*. **Hospital Regional No. 1 IMSS “Vicente Guerrero”, Acapulco, Guerrero:** *Josué Luis Medina Estrada MD. Intensive Care Unit*. **Hospital de la Beneficencia Española San Luis Potosí, San Luis Potosí:**

*Luis Arturo López Reveles MD, Emergency Department. Elsa Berenice Arriaga Rivera MD, Emergency Department*.

### Collaborators - monitors

Silvia Elena Uribe Moya MD – *Centro Medico Nacional IMSS “La raza”, Hospital de Infectologia “Dr. Daniel Mendez Hernandez”, Respiratory Care Unit*. Rodrigo Fernando Centeno Asencio MD **–** *Hospital General Regional Mérida “Ignacio García Tellez”, Emergency Department*. Alfredo García Tellez MD, Raymundo Montiel Latorre MD - *Hospital General de Pachuca; Emergency Department*. Luis Fernando Flores Zamora NUS - *Hospital Regional de la Universidad de Colima, Intensive Care Unit*. Dulce María Bernal Martínez MD, Victor Hugo García López MD - *Hospital General de Tláhuac, Mexico City*. Diego González Barbosa RT - *Centro Médico Nacional de Occidente, Guadalajara, Jalisco. Department of Respiratory Physiology*.

## Appendix 2. List of hospitals participating in the study and physicians in charge

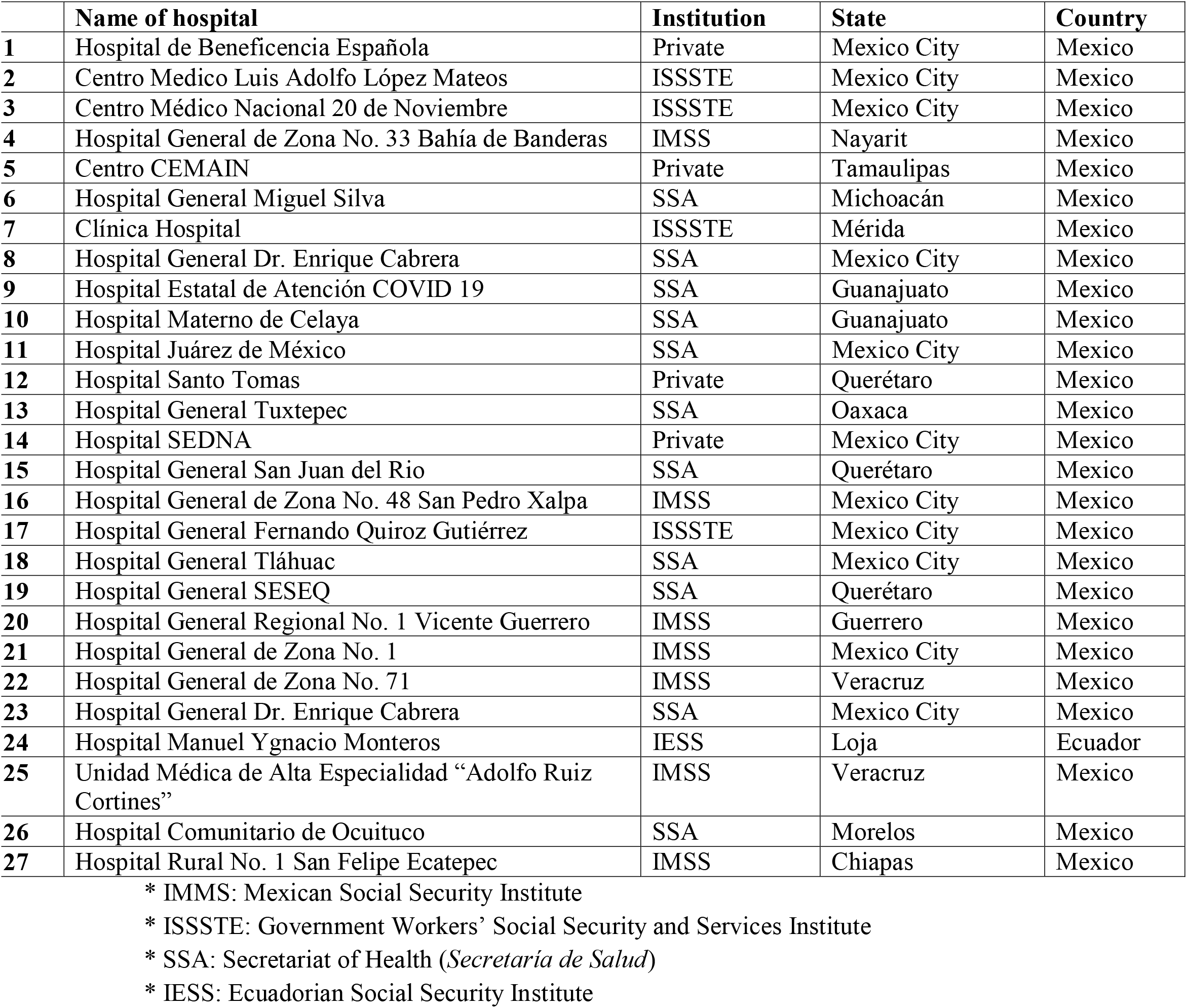

## Appendix 3. Chest CT assessment using the CO-RADS* categorical assessments scheme to evaluate suspicion of COVID-19

**Appendix 3.** Chest CT assessment using the CO-RADS\* categorical assessments scheme to evaluate suspicion of COVID-19
| **Category** | **Level of COVID-19 suspicion** | **Chest CT findings** |
| --- | --- | --- |
| **CO-RADS 1** | Very low | Normal or non-infectious abnormalities |
| **CO-RADS 2** | Low | Abnormalities consistent with infections other than COVID-19 |
| **CO-RADS 3** | Indeterminate | Unclear whether COVID-19 is present |
| **CO-RADS 4** | High | Abnormalities suspicious for COVID-19 |
| **CO-RADS 5** | Very high | Typical COVID-19 |
| **CO-RADS 6** | Proven | RT-PCR + for SARS-CoV-2 |

\*CO-RADS: COVID-19 Reporting and Data System.

## Appendix 4. Calculation of FiO_2_ based on type of supplemental oxygen delivery device used

**Appendix 4.** Calculation of $\text{FiO}_2$ based on type of supplemental oxygen delivery device used.
| Oxygen therapy | Flow (L/min) | *$\text{FiO}_2$ (%) |
| --- | --- | --- |
| Nasal cannula | 1 | 24 % |
|  | 2 | 28 % |
|  | 3 | 32 % |
|  | 4 | 36 % |
|  | 5 | 40 % |
|  | 6 | 44 % |
| Non-rebreather mask | 10-15 | 80-95 % |
| High-flow nasal cannula | Flows up to 60 | *Up to 100% |

\* $\text{FiO}_2$ : Fraction of inspired oxygen.

## Appendix 5. Density functions before and after propensity score matching of patients in the awake prone and awake supine cohorts

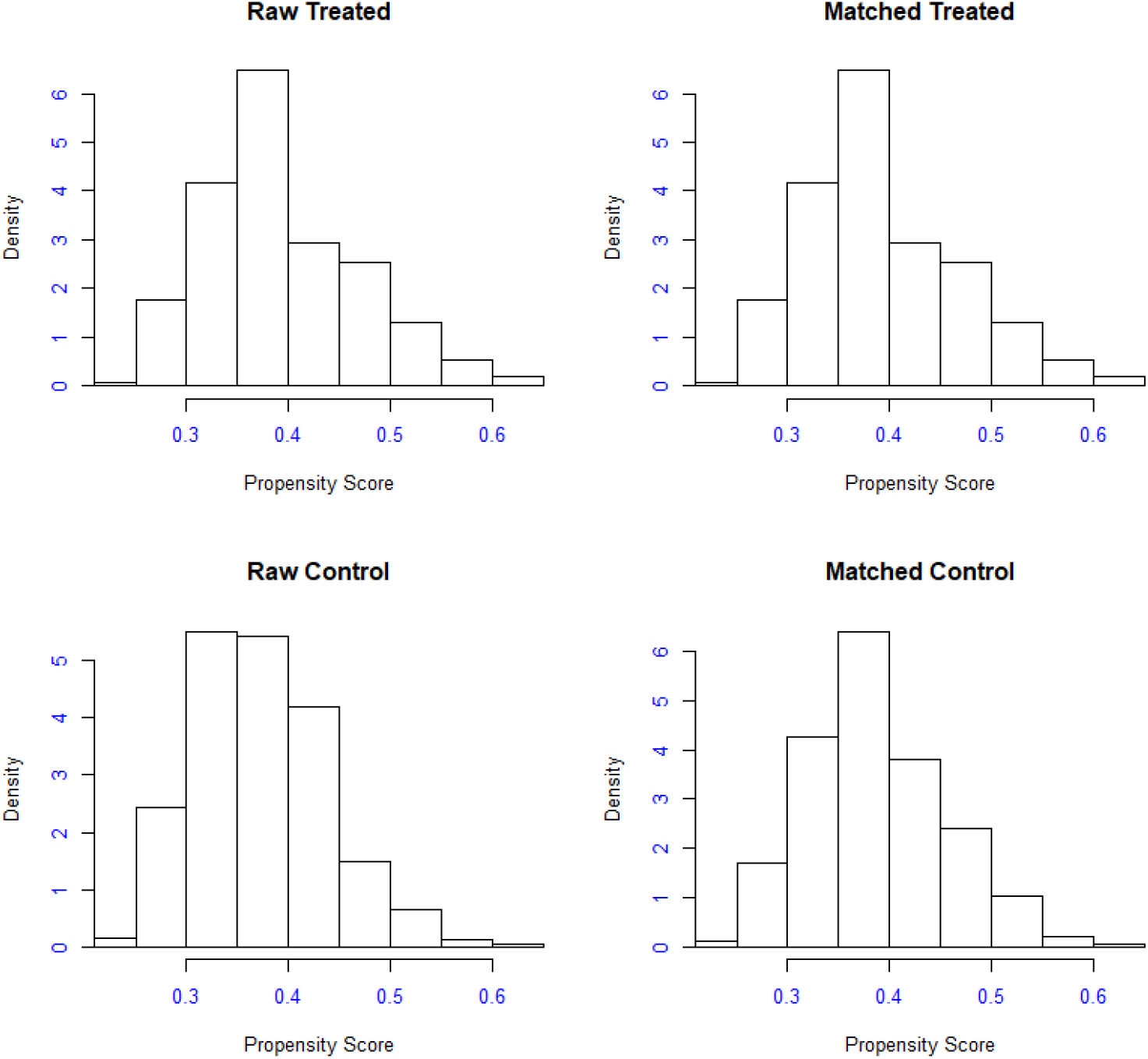

## Appendix 6. Search strategy

We searched MEDLINE and EMBASE through OVID, PubMed, BioRxiv and MedRxiv for research on COVID-19 published until 20 January 2021. We used the publicly available COVID-19 Living Evidence on COVID-19 dataset (31). Search terms for the search strategy were: (‘severe acute respiratory syndrome coronavirus 2’ [supplementary concept] OR ‘COVID-19’ [supplementary concept] OR ‘coronavirus’ OR ‘HCoV’ OR ‘nCoV’ OR ‘2019 nCoV’ OR ‘covid’ OR ‘covid19’ OR ‘severe acute respiratory syndrome coronavirus 2’ OR ‘SARS-CoV-2’ OR ‘SARS-CoV 2’ OR ‘SARS coronavirus 2’) AND (prone) AND (awake). The following filters were applied for study design: case series, case-control study, cohort study, trial, other, or unclassified. Studies were chosen regardless of language, provided an abstract in English was available, and if the study included and clearly differentiated patients undergoing awake prone positioning from those in awake supine position, as well as intubation rates for both groups.

## Appendix 7. Main characteristics of the classification and regression tree (CART) model

|  |  |
| --- | --- |
| Growing method | QUEST † |
| Dependent variable | Intubation |
| Independent variables | Sex<br>Age<br>Diabetes<br>Initial SpO<sub>2</sub>/FiO<sub>2</sub> ratio ††<br>Low-flow nasal cannula<br>High-flow nasal cannula<br>Non-rebreather mask<br>Prone position<br>Systemic arterial hypertension<br>Obesity<br>Heart disease<br>Lung disease<br>Cancer<br>Liver disease<br>Chronic kidney disease |
| Validation | Cross-validation |
| Maximum depth of trees | 5 |
| Minimum cases in parent node | 100 |
| Minimum cases in child nodes | 50 |
| Results |  |
| Independent variables included | Initial SpO<sub>2</sub>/FiO<sub>2</sub> ratio<br>Low-flow nasal cannula<br>Non-rebreather mask<br>Obesity<br>Diabetes<br>High-flow nasal cannula<br>Prone position<br>Cancer<br>Liver disease |
| Number of nodes | 5 |
| Number of terminal nodes | 3 |
| Depth | 2 |
| † QUEST: Quick, Unbiased, Efficient Statistical Tree. |  |
| †† SpO<sub>2</sub>; partial oxygen saturation, FiO<sub>2</sub> fraction of inspired oxygen. |  |

## Abbreviations

PP: prone position/prone positioning
PaO_2_: partial arterial pressure of oxygen
SpO_2_: peripheral arterial oxygen saturation
PaO_2_/FiO_2_: arterial partial pressure of oxygen /fraction of inspired oxygen
PaCO_2_: arterial partial pressure of carbon dioxide
RR: respiratory rate
NIV: non-invasive ventilation
HFNC: high-flow nasal cannula
ARDS: Acute respiratory distress syndrome
COVID-19: coronavirus disease 2019
STROBE: Strengthening the Reporting of Observational studies in Epidemiology
AP: awake prone
AS: awake supine
CO-RADS: COVID-19 Reporting and Data System
IQR: interquartile range
SD: standard deviation
CART: classification and regression tree
QUEST: quick, unbiased, efficient statistical tree
OR: odds ratio
CI: confidence interval
Ppl: pleural pressure
TPP: Transpulmonary pressure
V/Q: ventilation-perfusion.

